# The relationship between expelled eggs, morbidity and age: implications for current *Schistosoma mansoni* elimination policies

**DOI:** 10.1101/2024.12.04.24318477

**Authors:** Rivka M. Lim, Ruhi Lahoti, Moses Arinaitwe, Victor Anguajibi, Sergi Alonso, Andrina Nankasi, Fred Besigye, Alon AtuhAire, Amy B. Pedersen, Joanne P. Webster, Poppy H.L. Lamberton

## Abstract

Direct morbidity assessments are rarely included in monitoring and evaluation of *Schistosoma mansoni* control programmes. Instead, the number of eggs-per-gram (EPG) of faeces is used as a proxy for morbidity. The World Health Organization targets schistosomiasis elimination as a public health problem (EPHP) by 2030, defined as <1% heavy infections (≥400 EPG for intestinal schistosomiasis). However, recent findings challenge this link between infection intensity and morbidity.

Prevalence and intensities of *S. mansoni* infection were diagnosed by Kato-Katz and point-of-care circulating cathodic antigen tests in 287 individuals, 3-74 years-old, from Bugoto, Uganda. Ultrasound examinations following the Niamey protocol characterised metrics of morbidity: periportal fibrosis (PPF), portal vein dilation (PVD) and left parasternal line (PSL) enlargement. Additional morbidity markers included anaemia and self-reported symptoms. Malaria status was determined using rapid diagnostic testing. Logistic regression models elucidated potential predictors of morbidity.

PPF, PVD, PSL and anaemia prevalence were 9%, 34%, 33% and 13% respectively. School-aged children (SAC) had the highest infection intensity, but pre-school-aged children (PSAC) were significantly more likely to have PVD, PSL and anaemia than other age groups. Current *S. mansoni* infection predicted only self-reported symptoms of blood in stool and rash. As infection intensity increased, so did the likelihood of anaemia and fibrosis, but this was significant only at levels much higher than the 400 EPG threshold. Current malaria infection, measured with rapid diagnostic testing, was associated with PVD and anaemia.

Our findings add to growing evidence against using ≥400 EPG as a proxy for schistosomiasis morbidity within control programs, urging a revaluation of targets. The age-related distribution of morbidities observed, with a notable burden in PSAC, highlights a critical need to elucidate the impact of less-specific morbidities on host health and its interplay with current and past infections with *S. mansoni* and other parasites.

**Author Summary:** Schistosomiasis is a parasitic disease that affects millions of people worldwide, especially in sub-Saharan Africa. Monitoring and control programs typically rely on counting parasite eggs in stool as a proxy to measure disease severity, but recent evidence questions whether this method accurately reflects the real health impact of the disease.

Here we examined 287 individuals in Bugoto, Uganda, aged 3 to 74, using both stool and blood tests to detect *Schistosoma mansoni*infection. We also used ultrasound to assess liver damage and other health complications caused by the parasite and used rapid testing of participants blood to diagnose malaria. Surprisingly, we found that even at very low *S. mansoni* infection levels, young children had significant liver-related complications and anaemia. We also found that only very high egg counts were associated with severe health problems such as liver fibrosis. Additionally, current malaria infection was strongly linked to anaemia and liver abnormalities, complicating the picture further.

Our findings suggest that the current World Health Organization thresholds for defining disease severity does not provide an accurate picture of the true burden of schistosomiasis, particularly in young children and in regions of co-endemicity with malaria. This research highlights the urgent need for new methods to assess the impact of the disease.

## Introduction

Schistosomiasis is a parasitic disease caused by trematodes of the *Schistosoma* genus. It is a substantial cause of global morbidity, with the highest disease burden found within sub-Saharan Africa [1]. *Schistosoma mansoni* is one of the key species responsible for the hepato-splenic and intestinal forms of the disease in humans, and infection is acquired by contact with freshwater infested with cercariae larvae shed from intermediate *Biomphalariaspp* snail hosts. Cercariae penetrate skin and enter the bloodstream of a human host where they migrate through the heart and lungs to the liver, where they mature and find a mate [2,3]. Following this, most adult *S. mansoni* worms travel to and reside in the portal vein and mesenteric venules which drain the intestines, and adult worm pairs are estimated to produce up to 300 eggs per day (Gryseels and de Vlas, 1996). These laterally-spined eggs are either excreted from the body in the faeces, which contributes to onward transmission, or become trapped in host tissues causing immunopathological reactions and the major clinical manifestations associated with the disease [5]. Due to the direction of blood flow, a significant number of eggs are carried to the liver, here they become lodged in tissue causing inflammation, granulomas, damage and scarring, which can obstruct the veins surrounding the liver and cause hepatomegaly, portal hypertension, liver disfunction and periportal fibrosis (PPF) [6,7]. Infection can also cause broad symptoms such as abdominal pain, diarrhoea, and blood in the stool [8], as well as additional clinical features such as malnutrition, anaemia, stunting, and cognitive dysfunction [7], reduced physical fitness and aerobic capacity [9].

*Schistosoma mansoni* is most commonly diagnosed by visualising eggs expelled in the faeces using light microscopy and the Kato-Katz thick smear method (Kato-Katz) [10]. More recently, the World Health Organization (WHO) have also endorsed the use of point-of-care circulating cathodic antigen (POC-CCA) tests for *S. mansoni* diagnosis. These tests detect antigens regurgitated by feeding adult worms, which are excreted in urine [11]. POC-CCA tests have higher sensitivity than Kato-Katz in detecting low-intensity infections [12], however without well-defined indications of how POC-CCA aligns with infection intensity, the WHO currently only recommend this test for prevalence diagnostics rather than infection intensity [13].

Clinical manifestations of *S. mansoni*-associated morbidity can be diagnosed using various methods. The Niamey Protocol was developed to standardise the assessment of *Schistosoma*-associated morbidity using ultrasound examination. This protocol primarily focuses on detecting organ dysfunction, PPF, portal hypertension, hepatosplenomegaly and ascites [14]. Other diagnostic methods, which are often less specific, include, faecal occult blood point of care tests, visual identification of blood in stool, diarrhoea, self-reported abdominal pain and blood tests for anaemia [15]. Morbidity data are, however, generally not collected during routine Mass Drug Administration (MDA) surveillance, monitoring and evaluation activities. Instead, current faecal egg counts are used as a proxy for morbidity with those shedding a high number of eggs assumed most at risk for morbidity [16–18]. Eggs per gram (EPG) of faeces is calculated from Kato-Katz to determine the intensity of infection of intestinal schistosomiasis, with light, moderate and heavy intensity infections classified as 1-99, 100-399 and ≥400 EPG, respectively [19].

The main method of schistosomiasis control recommended by the WHO is MDA, in which school-aged children (SAC, 6–14-year-olds) and at-risk adults are treated with the anthelminthic drug praziquantel, regardless of individual current infection status [20,21]. This is effective at reducing infection intensities and killing adult worms, however, praziquantel is not effective on juvenile worms, does not prevent reinfection, and disease symptoms and associated morbidity may not be reversable with treatment especially if the infection is chronic or severe [22–24]. Chronic morbidity associated with schistosomiasis is assumed to develop over time, with more severe liver fibrosis and other enduring conditions observed in older-aged populations residing in endemic regions, likely due to cumulative infections [25–27]. The initial objective of MDA programmes was to reduce morbidity by reducing infection intensities [20,28], with treatment focussed on SAC to reduce infections before severe morbidity had established and prevent chronic pathologies from developing as these individuals age. Using a morbidity threshold set by the WHO, if there are <5% of heavy intensity infections in SAC, a community was considered to have morbidity under control [18]. However, there is growing recognition that morbidity is occurring in both Pre-SAC (PSAC) [29,30] and SAC, not only adults. Therefore, in the most recent WHO Neglected Tropical Disease Roadmap the aim is to mitigate the impact on child health, with the recommendation to include MDA for all children over two years of age [13], and this is to be facilitated by the development of a new paediatric formulation of praziquantel [31]. This new roadmap also has revised targets for schistosomiasis control, aiming for elimination as a public health problem (EPHP) in all endemic states by 2030 [32] with an EPHP threshold of <1% heavy intensity infections [13]. Consequently, the relationship between intensity of infection and morbidity is integral to effective disease control.

Research is emerging, however, which challenges the use of current *S. mansoni* egg shedding as predictive of morbidity. One recent cohort study of SAC across six African countries collected data using ultrasound, clinical findings, and self-reported symptoms and observed no consistent association between these morbidity markers and current infection intensity category, when these are grouped by the WHO intensity categories for *S. mansoni* [33]. An earlier comparative study from Egypt and Kenya found no clear epidemiological association between egg counts (EPGs) and morbidity markers such as PPF or portal hypertension, despite substantial differences in the geometric mean EPG and the prevalence of these conditions between the two regions [27]. In addition, current thresholds may not adequately take into consideration the differences between heavy and moderate infection intensities [34,35], or account for the impact of decades of MDAs, that may have mitigated disease progression, leaving individuals with underlying morbidity or undetectable infections which could be missed by current guidelines. This possible lack of a relationship between egg count and morbidity suggests there is a need for more up to date information on age-specific morbidity markers in monitoring and evaluation of *S. mansoni* control programmes. With large scale schistosomiasis control and elimination programs now more common place, the increase to biannual treatment and additional interventions in high endemic regions, some researchers are suggesting there is going to be a shift from the more severe cases of PPF to more subtle morbidities [35]. This raises the question of whether these low-intensity infections, or a negative Kato-Katz test but a positive POC-CCA should be used to identify subtle morbidities and prevent progression to more chronic and less treatable conditions [34]. A suggestion has been made to redefine the current definition of EPHP and morbidity control for schistosomiasis using overall prevalence of schistosome infection rather than heavy intensity infections only [35,36].

A further complication is the effects of other diseases or comorbidities. PPF as diagnosed by the Niamey Protocol is aimed to be specific to *S. mansoni* associated fibrosis. However, less specific tests such as liver and spleen enlargement, may complicate the accurate designation of morbidity causes, particularly in regions co-endemic with other infectious diseases such as malaria and soil-transmitted helminths (STHs), which can present with similar morbidity outcomes [29].

Disentangling the drivers of morbidity across different age groups remains a challenge for how best to optimise resource allocation and treatment strategies. Whilst previous studies have largely focused on SAC, there is a notable gap in research that examines morbidity and the influence of co-endemic infections on morbidity across the full spectrum of age groups in the same community, especially for PSAC. Additionally, although both the Kato-Katz method and the more sensitive POC-CCA diagnostic have been employed in past studies, there is a lack of research that systematically applies these diagnostics alongside multiple morbidity predictors—both specific and nonspecific—across all age groups.

Therefore, the main objectives of this study were to characterise *S. mansoni* prevalence, infection intensity, and liver morbidity within a population residing in an area known for high *S. mansoni* endemicity. Specifically, we aimed to determine whether adults exhibit a greater burden of liver morbidity compared to children (PSAC and SAC), potentially reflecting the impacts of long-term chronic exposure. Additionally, we sought to examine the relationship between *Schistosoma*-associated morbidity and the presence of current infections, including *S. mansoni*, malaria, or STHs. A further goal was to assess the power of *S. mansoni* infection intensity in predicting *Schistosoma*-related morbidity and to explore associations with other potential predictors of morbidity, such as malaria, STH infection, and age. Finally, the study aimed to elucidate if individuals who are *Schistosoma*-negative in this highly endemic population are more likely to exhibit liver morbidity, possibly due to previous exposures and infections, when compared to uninfected healthy controls from non-endemic areas who were instrumental in developing the Niamey protocol.

## Methods

### Ethical Statement

Ethical clearance was granted by the Uganda National Council for Science and Technological Social Sciences (reference: UNCSTHS 2193), the Ugandan Ministry of Health Vector Control Division Research Ethics Committee (reference: VCDREC/062) and the University of Glasgow College of Medical, Veterinary and Life Sciences (reference: MVLS tel:200160068). Informed consent was gathered prior to any data collection, from all participants over the age of 18 by either signature or thumb print, whilst parents or legal guardians gave consent for those under 18 years old, and informed assent was obtained from all children eight to 18 years old. All participants were made aware that they were allowed to opt out of the study at any point without it affecting their praziquantel treatment from this study or from the national control programme.

### Study area

This study was carried out in May 2022 in Bugoto community, which comprises of two villages: Bugoto A and Bugoto B, located on the north shores of Lake Victoria, Mayuge District, Eastern Uganda. Lake Victoria is well known for being highly endemic for *S. mansoni* [37–39] and these small rural village communities are mostly involved in small scale agriculture and fishing, where daily interactions with the lake are frequent [37]. Annual MDA with praziquantel has been taking place in these villages to all SAC since 2003, then community-wide (SAC and adults only, not including PSAC) since 2004 and then in 2019 this increased to community-wide biannual (SAC and adults only still) treatment [28,37,40]. This area is also highly endemic for malaria infection caused by *Plasmodium falciparum*, with a prevalence of 21% in children <5 years old in the sub-region of Busoga [41,42]. Malaria control measures include community-wide distribution of insecticide-treated nets every three years, health-centre-based treatment for those seeking it and integrated community case management (ICCM) of malaria in children, which is test and treat for suspected malaria patients [41]. Annual praziquantel MDAs also include treatment of 400mg albendazole for soil transmitted helminths, which are often co-endemic [38].

### Data Collection

Data were gathered over a two-week period in May 2022, in the wet season and just prior to the annual national MDA, when people are expected to have the highest infection intensities. All members of the village apart from pregnant women and children under 2 years old were eligible to participate and Village Health Team members assisted with recruitment and informed consent, approaching 400 participants stratified across ages with an equal sex ratio. Parasitological, clinical and ultrasound measurements were performed in the field. A bespoke questionnaire was used to retrieve information on the height, weight, age and sex of each participant along with questions about schistosomiasis-related symptoms they had experienced in the last month, previously described in full [43]. Smartphones using ODK software were used to collect data.

Three stool samples from each participant were obtained on three separate days and duplicate Kato-Katz [10] were prepared per stool. These smears were examined within one hour of preparation to quantify *S. mansoni , Ascaris lumbricoide*, *Trichuris trichiura*, *Enterobius vermicularis, Hymenolepis nana* and hookworm eggs. Individual infection, infection intensity (EPGs) and community infection prevalence were calculated. Urine samples were collected for POC-CCA cassette testing (Schisto POC-CCA®, ICT International, Cape Town, RSA) following the manufacturer’s instructions, and semiquantitative results (G1 to G10) were assigned using the G-score system [44]. An individual was considered positive for *S. mansoni* if they were either egg positive by Kato-Katz or egg negative by Kato-Katz but had a POC-CCA G-score of ≥3 [45]. Malaria infection was determined using finger prick blood (Ag P.f/Pan Malaria Rapid Diagnostic test, Standard Diagnostics Inc.) [46], which can diagnose *Plasmodium* species in whole blood. At the end of the study and in accordance with national guidelines, all participants were given a snack and offered praziquantel at 40mg/kg, 400mg albendazole and those diagnosed as positive for malaria, were offered dihydroartemisinin/piperaquine, the current WHO recommended treatment, given directly to them or the parents/legal guardians of those diagnosed with malaria. Any participant observed to have severe pathology were referred to the appropriate health authority.

### Ultrasound examination

Each participant was examined in the supine position using the portable ultrasound machine, Aloka Prosound 2 (Hellige, Freiburg, Germany), equipped with curve-linear probe and variable frequency of 2.5-6MHz. These were performed by a single senior radiographer trained in the Niamey protocol [14] from the Ministry of Health in Uganda who was blinded to the infection status of the individual. The left liver lobe size was measured from the upper to the caudal margin in the left parasternal line (PSL) of the lobe to indicate for left liver lobe enlargement. Portal Vein Diameter (PVD) measurements and the presence of any ascites or collaterals was also recorded and were taken to gauge for risk of portal hypertension. Both the PSL and PVD measurements were standardised by height and compared to healthy Sudanese controls from a non-endemic population [14,47]. Liver parenchyma patterns were logged according to the protocol to determine degree of PPF. Each liver image pattern (IP) was designated a score from A-F, where A is classed as no morbidity, B is undetermined and C-F (and combinations therefore of are scored). The spleen was not measured as per instructions in the protocol for regions which are endemic for malaria, however the sonographer did note if observed splenomegaly.

### Data processing

Data were exported to R version 4.2.2 where all data management and statistical analysis were executed. Seven age groups were established from the study population for descriptive analysis: i) 3 – 5 years, ii) 6 – 10 years, iii) 11 – 14 years iv) 15 – 20 years, v) 21 –30 years, vi) 31 – 40 years, and vii) > 40years. Three age classes were also established for statistical modelling: i) 3-5 years = PSAC, ii) 6-15 years = SAC, and iii) >15 years = adults.

To score morbidity in the participants, we used a binary system. To measure PPF, a score of 0 was given if the ultrasound designated IP of A or had either X, Y or Z pattern scores as these are deemed non-*Schistosoma* related morbidities [14] and a score of 1 if B-F or combinations thereof. As we did not have data on the thickness of the walls and branches of the portal vein, we did not calculate a periportal thickening (PT score) as per Niamey protocol. PVD was determined by comparing the PVD to height-matched healthy controls [14,47], a PVD score was given depending on if the PVD was normal size (score = 0) or dilated in which it was >2 standard deviations (SD) above the healthy comparisons (score = 1). The left liver lobe measurement was also compared to healthy standards [14,47] if not enlarged (score = 0) or enlarged in which it was >2 SD larger than healthy comparators (score = 1). Anaemia was categorised as either positive (score = 1) or negative (score = 0) for logistic regression modelling or normal, mild, moderate and severe when used as a descriptor, calculated as referenced by the WHO for Hb (g/l) measures at 1200 metres above sea level [48]. A score of 1 was given each time a participant answered yes to survey questions regarding symptoms which could be attributed to *S. mansoni* infection: diarrhoea, blood in stool, abdominal pain, pain when urinating, nausea, vomiting, headache, fever, dizziness, body swelling, chills, difficulty breathing, rash and/or weakness.

### Statistical analysis

#### Characterise S. mansoni prevalence, intensity, and liver morbidity within a highly endemic population

Patterns in *S. mansoni* infection intensity across ages and sexes were analysed through a general linear model (GLM), stratified by age group and sex, of the arithmetic mean of infection intensity for each individual. The Shapiro-Wilk test of normality was used to assess whether the distribution of individuals negative by Kato-Katz but positive by POC-CCA in different age groups deviated from a normal distribution and skewed towards one age group. To characterise morbidity across ages and sexes, logistic regression models were used with morbidity as a binary response variable for each morbidity marker.

For malaria infection characterisation, a logistic regression GLM was used with malaria infection as a binary response variable and age group and sex as predictors. Due to the 6–10-year-olds having the highest overall prevalence of malaria, this group was used as reference in the model.

#### To determine if adults have higher liver morbidity relative to children (PSAC and SAC), as predicted from longer term chronic S. mansoni exposure

To elucidate the relationship between non-specific liver morbidity and age, a logistic regression model was conducted with morbidity as the binary response variable (see data processing) and including age as both a linear and quadratic function. The vertex of the quadratic function was calculated and used to infer the minimum or maximum age at which the relationship changed.

#### Evaluate the association between morbidity and a current S. mansoni, malaria or STH infection

Univariate logistic regression models were carried out with each of the morbidity markers as response variables. The predictors were *S. mansoni* as diagnosed by Kato-Katz only, *S. mansoni* infection as diagnosed as a combination of Kato-Katz and CCA ≥3, age class, sex, malaria status and hookworm infection status as categorical variables.

#### Evaluate if S. mansoni infection intensity predicts morbidity and test associations with other potential predictors of morbidity

Multivariate logistic regression models were caried out using only individuals who were positive for *S. mansoni*via both diagnostic techniques (Kato-Katz positive or Kato-Katz negative and CCA ≥3 positive), with each of the morbidity markers as response variables. Given that malaria can cause anaemia, PVD and PSL, and that we observed age trends in the descriptive analysis of these morbidity markers; age class and current malaria infection were added along with log mean intensity of *S. mansoni* infection as predictors in the models for these response variables. When PPF was the response variable, the model did not include age class as a predictor due to the very low number of fibrosis cases in each age class, including this would have risked class imbalance. If a positive significant relationship was estimated between log mean intensity of infection and a morbidity marker, the model was rerun whilst sequentially removing the highest infection intensity points to determine the intensity level which was driving the significant relationship.

#### To determine if S. mansoni negative individuals from this highly endemic population are more likely to have liver morbidity than the uninfected healthy controls used to develop the Niamey protocol who are from a S. mansoni non-endemic area

Since the raw data for healthy comparisons of PVD and PSL were not available (Yazdanpanah et al., 1997 – author contacted), a dataset was simulated using the graph and standard deviations provided in Annex C: Organometry of the Niamey Protocol [14]. These data were generated by resampling 1000 bootstraps and using the mean and standard deviations from the protocol and matching the sample sizes per height category found in our dataset. Summary statistics were calculated and compared. A two-sample Welch t-test was conducted to compare the mean measurements for each height category between the simulated data and the study data.

With all GLMs, unless otherwise stated, the reference groups for categorical variables were as follows: age group = 11-14 (as this had the highest *S. mansoni* prevalence and intensity of infection), age class = adults, sex =female and for all binary variables a negative outcome was used as reference.

## Results

### Bugoto is a S. mansoni high endemicity region with low periportal fibrosis and moderate unspecific liver morbidity

Participants who had complete data for sex, age, PVD, IP, PSL scores and at least one Kato-Katz sample were included in the analysis, totalling 287 participants: including 104 male and 188 female participants with an age range of 3-74 years, and a median age of 14. Three stool samples were collected for 192 (67%) participants, 89 (31%) people provided two samples, and 6 (2%) people provided only one stool sample. Three days of urine samples were collected for 257 individuals (90%), 26 (9%) with samples over two days and 4 (1%) with only one urine sample.

Data from the study participants supports the categorisation of these Bugoto communities as being highly endemic for *S. mansoni* with an 81% prevalence in SAC, as measured by just one Kato-Katz, as per the WHO guidelines, which rises to 91% infection prevalence measured by composite reference including up to six Kato-Katz and POC-CCA G-score ≥3 (Figure 1A - black). When measured for egg positivity by Kato-Katz only, community prevalence was 58% (Figure 1A - blue), however 43 individuals were egg negative by Kato-Katz but positive by POC-CCA with a G-score of ≥3. This was disproportionally found in the younger age groups, with 11 samples from the PSAC being negative by Kato-Katz but positive by POC-CCA (skewness=0.69, Shapiro-Wilk test for normality, W=0.89, p=<0.001) (Figure S1). Arithmetic mean egg intensity of the whole community was 139 EPG (range: 0-2136) (Figure 1B). Both prevalence and intensity of infection peaked in 11–14-year-olds, with males in this age category having a significantly higher infection intensity than females (mean difference: 1.148, p=0.013) (Figure S2). An EPG of ≥400 (heavy infection according to WHO) was found in 11% (n=32) of participants (Figure 2B).

**Figure 1.**
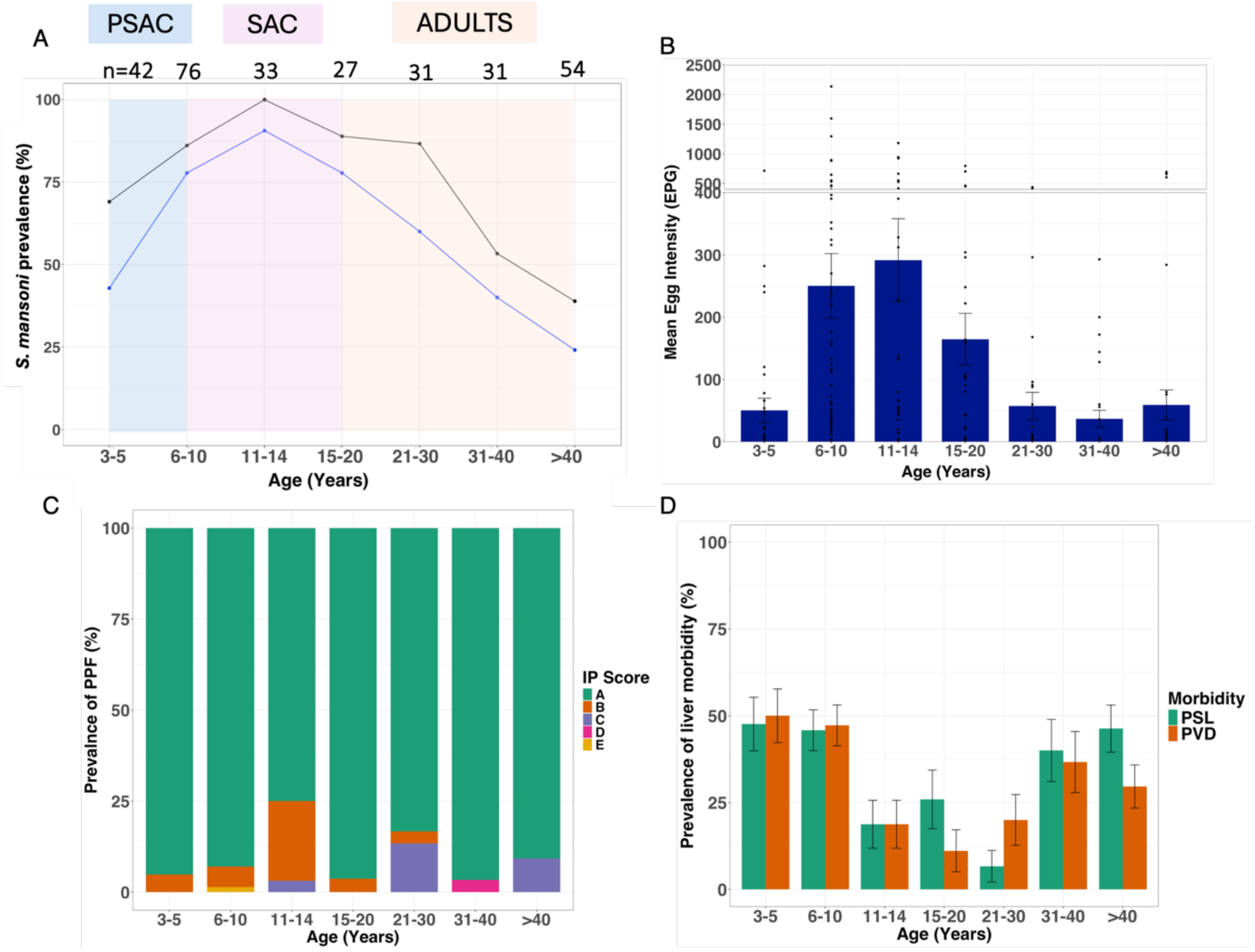
Characterisation of <u>Schistosoma mansoni</u> by age in Bugoto, Uganda in 2022. (A) Community prevalence as measured by Kato-Katz (blue) and Kato-Katz plus point-of-care circulating cathode antigen (POC-CCA) G-score ≥3 (black). Shaded areas are indicating preschool agechildren (PSAC) = blue, school age children (SAC) = pink and adults = orange. (B) Mean egg intensity as measured by eggs per gram (epg) in faeces, with the separate section at the top indicating those with heavy infections (≥400 epg). (C) Prevalence of individuals with liver image pattern (IP) scores A-E, and (D) prevalence of unspecific liver morbidity as measured by portal vein diameter (PVD) and parasternal line of the left liver lobe (PSL). Error bars are standard error of the mean (B), and binary standard errors.

**Figure 2:**
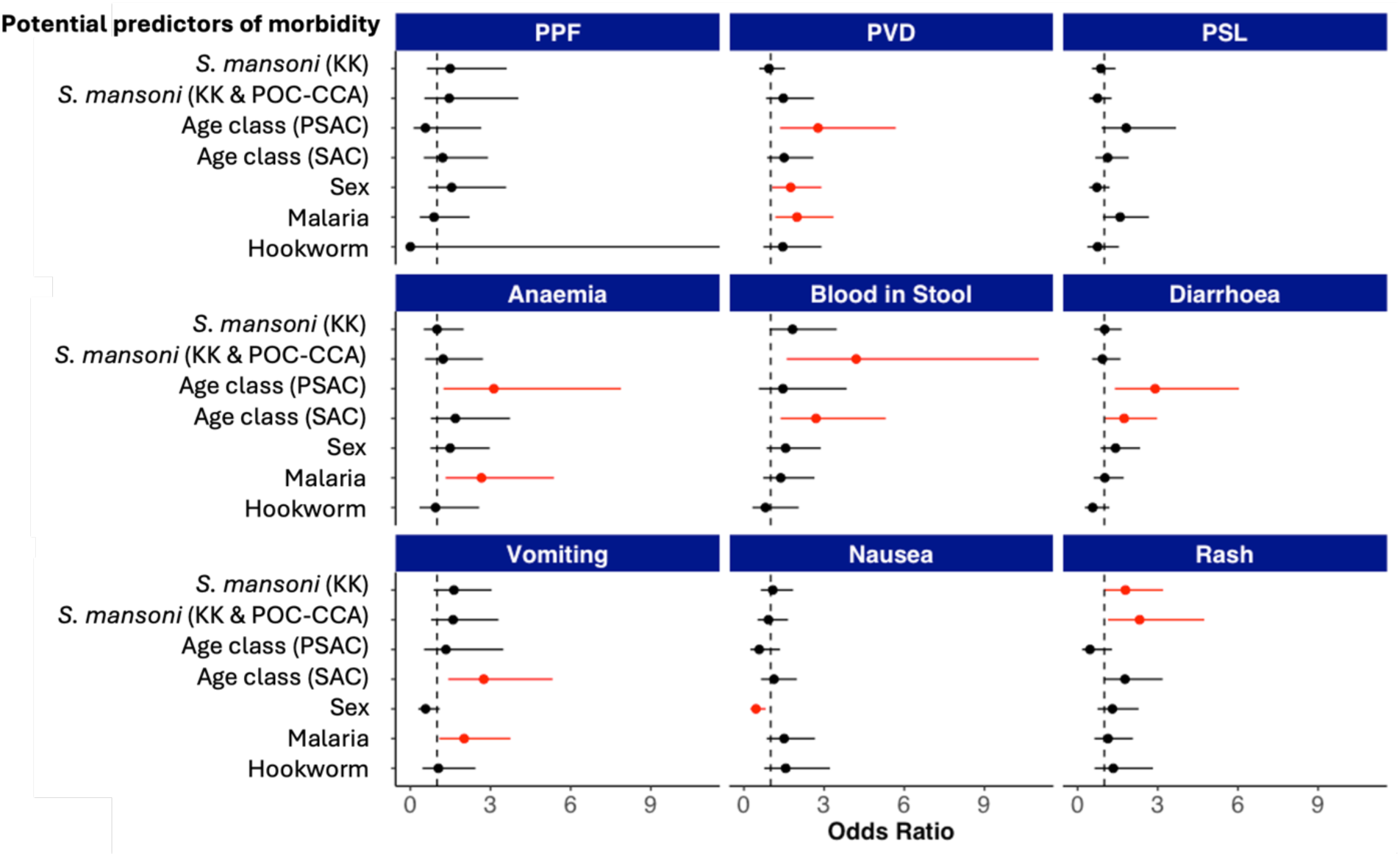
Current <u>Schistosoma mansoni</u> infection as a predictor of morbidity. Univariate logistic regression models with periportal fibrosis (PPF) as measured by IP score, portal vein diameter (PVD), left liver lobe enlargement as measured by parasternal line (PSL), anaemia and blood in stool, diarrhoea, vomiting, nausea and rash as binary response variables. Predictors areS. mansoni as measured by Kato-Katz (KK) only, by KK and point of care circulating cathode antigen (POC-CCA) ≥3, age classes are preschool age children (PSAC) and school age children (SAC) with adults as the reference group, the reference group for sex was female. Each predictor has a point estimate of odds ratio as predicted from the model and 95% confidence interval. Vertical dashed line indicates odds ratio = 1, no effect. Red point estimate is significant if p-value = <0.05.

Stool samples examined for non-*S. mansoni* parasite eggs were available for 285 participants. Hookworm infection was identified in 14% of the population, *A. lumbricoides* 0.3%, *H. nana* 0.3%, *E. vermicularis* 0.6% and *T. trichiura* 0.6%. There was no significant difference in prevalence of hookworm by age classes nor did the likelihood of hookworm infection increase with age. A positive malaria test was observed in 31% of the community (n=282). Among age groups, 6–10-year-olds exhibited the highest prevalence of malaria at 60% (42 out of 70), and were significantly more likely to be infected with malaria with an odds ratio of 1.50 compared to other age groups: 3-5 years (OR=0.31, p=0.005), 11-14 years (OR=0.32, p=0.012), 15-20 years (OR=0.28, p=0.009), 21-30 years (OR=0.07, p<0.001), 31-40 years (OR=0.17, p<0.001), and >40 years (OR=0.10, p<0.001) (Figure S3). Coinfection with malaria, hookworm and *S. mansoni* was observed in 5% of the population, while coinfection of just *S. mansoni*and malaria was more common (26%; 74 out of 282), with the majority of these cases found in the 6–10-year-olds (53%).

The most common self-reported symptoms in all age classes were headache (73%), abdominal pain (64%), diarrhoea (47%) and fever (44%), whilst the lowest reported were body swelling (8%) and difficulty breathing (7%). School age children were significantly more likely to report blood in stool than other age classes (OR=2.84, p=0.002) and PSAC and SAC were significantly more likely to report diarrhoea than adults (ORs=2.79 and 1.85, p=0.006 and 0.025 respectively). SAC and adults were significantly more likely than PSAC to report being dizzy, having a rash, vomiting or weakness (Table 1).

**Table 1.**
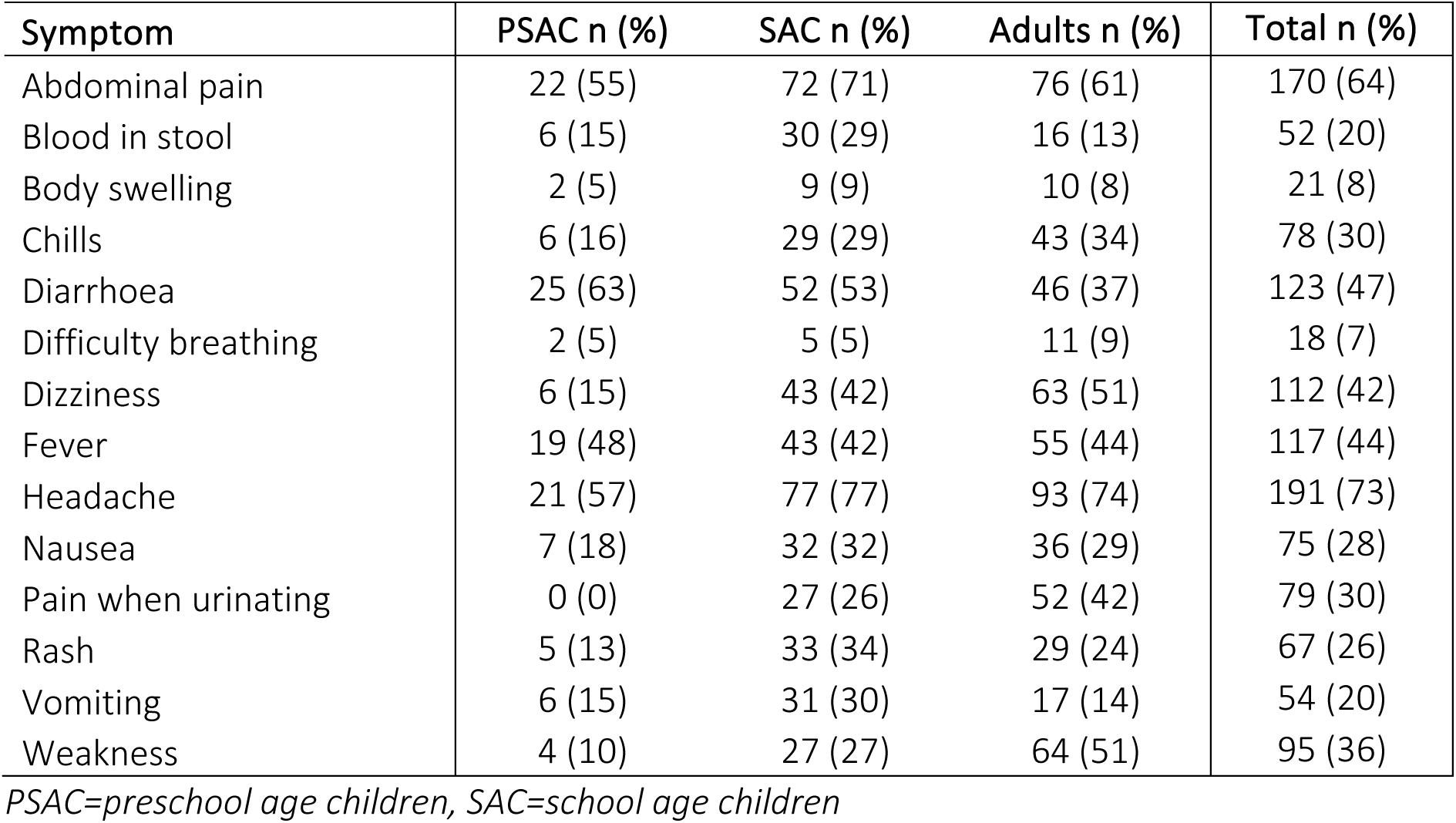
Number and frequency of participants who answered yes to having a self-reported symptom in the last month.

| Symptom | PSAC n (%) | SAC n (%) | Adults n (%) | Total n (%) |
| --- | --- | --- | --- | --- |
| Abdominal pain | 22 (55) | 72 (71) | 76 (61) | 170 (64) |
| Blood in stool | 6 (15) | 30 (29) | 16 (13) | 52 (20) |
| Body swelling | 2 (5) | 9 (9) | 10 (8) | 21 (8) |
| Chills | 6 (16) | 29 (29) | 43 (34) | 78 (30) |
| Diarrhoea | 25 (63) | 52 (53) | 46 (37) | 123 (47) |
| Difficulty breathing | 2 (5) | 5 (5) | 11 (9) | 18 (7) |
| Dizziness | 6 (15) | 43 (42) | 63 (51) | 112 (42) |
| Fever | 19 (48) | 43 (42) | 55 (44) | 117 (44) |
| Headache | 21 (57) | 77 (77) | 93 (74) | 191 (73) |
| Nausea | 7 (18) | 32 (32) | 36 (29) | 75 (28) |
| Pain when urinating | 0 (0) | 27 (26) | 52 (42) | 79 (30) |
| Rash | 5 (13) | 33 (34) | 29 (24) | 67 (26) |
| Vomiting | 6 (15) | 31 (30) | 17 (14) | 54 (20) |
| Weakness | 4 (10) | 27 (27) | 64 (51) | 95 (36) |

PSAC=preschool age children, SAC=school age children

Overall, 13% of the population had a positive anaemia diagnosis (n=287). Anaemia was highest 24% (10 out of 42) in PSAC, followed by 13% in SAC and 9% in adults (Table S1). Of these positive diagnoses, more than half were mild anaemia (8% of the population), 5% exhibited moderate anaemia, while notably, two individuals presented with severe anaemia. The first case involved a 38-year-old female who tested negative for all parasites screened but showed evidence of enlarged PVD and PSL. She reported 12 out of 14 clinical symptoms over the past month, and the sonographer reported gross splenomegaly secondary to portal hypertension. The second case was an 8-year-old male who tested negative for malaria, hookworm and other helminths, *S. mansoni* by Kato-Katz but positive with a POC-CCA test for schistosomiasis. He also had PVD and PSL enlargement and reported symptoms of blood in stool, diarrhoea, abdominal pain, and headaches over the past month.

*Schistosoma mansoni-*specific morbidity, as measured by liver image pattern (IP) scores, was not common in this community. Out of the 287 examined by ultrasound, a score of B-F was observed in just 27 participants from the ages of 4-74 years (9%) and 12 with a positive IP score of C-F (4%) (Figure 1C). A score which could be deemed as non-schistosomiasis associated ( X, Y, Z in the grading system) were found in seven participants and they were allocated zero for the IP (X (cirrhosis)=0, Y (fatty liver)=6 and Z (course liver)=1).

A total of 97 people (34%) had a PVD of at least two standard deviations above the Niamey Protocol healthy comparators [14,47]. An increased PSL of the left liver lobe, with a score of at least 2 standard deviations above healthy comparators were found in 94 (33%) participants. The incidence of both PVD and PSL overlapped in individuals with the two morbidity profiles being highly correlated (X^2^=35.5, p<0.001) and 50% of the population had either increased PSL, increased PVD or both (Figure 1D). No participants were observed to have either collaterals or ascites during ultrasound examination.

### Indeterminate fibrosis (IP=B) and unspecific liver morbidity were higher in the younger age groups than in adults in Bugoto, Uganda

In Bugoto, adults (>15 years of age) were not found to carry a greater burden of PPF when this measurement includes an IP score of B (indeterminate fibrosis) compared to PSAC (IP-BF: OR=0.53, p=0.431) or SAC (IP-BF: OR=1.18, p=0.712). The age group with the highest intensity of infection (11–14-year-olds) were significantly more likely to have a positive B-F score than all other age groups. However, this result is driven by the presence of IP B (n=15) rather than IP scores which represent pronounced fibrosis (C-F). IP B on its own was highest in SAC (n=11), and only two cases in each PSAC and adult age groups (Figure 1C).

Apart from one 8-year-old girl with an IP score of E and mean EPG of 2150 (Figure 1B), all cases of IP scores C and D (n=11) were observed in the older age groups. These scores were absent from PSAC and younger SAC (6–10-year-olds), with only one case in 11–14-year-olds, and most prevalent in the 21–30-year-olds (13%) and >40’s (9%). An IP score of D, or its combinations, was observed only once in the population and this was in the 31-40-year-old group. Therefore, the trend of more pronounced fibrosis (C-D) appears skewed towards adults; however, the small number of people with pronounced fibrosis in our sample precludes meaningful statistical testing. No community member was observed with an IP score of F or combinations which include F.

Unspecific liver morbidity as measured by a combination of PVD and PSL was more common in both the older and younger age groups, with intervening years (11–30-year-olds) carrying a lower burden (Figure 1D). PSAC were more likely to score positive for unspecific liver morbidity (PVD + PSL) than adults (OR=2.47, p=0.015). When examining the shape of the relationship between age and unspecific liver morbidity, we observed that once an individual is past PSAC age, the risk of unspecified liver conditions decreases with increasing age. However, a positive quadratic term for age indicates that this decreasing trend reverses at approximately 32 years of age, after which the risk of liver morbidity begins to increase, though it does not reach the high levels observed in PSAC (age as linear term (b): est= -0.0867430, p<0.001; age as quadratic term (a): est=0.0013662, p=0.001; vertex=b / (2* a)).

### Current S. mansoni infection predicts blood in stool and rash but no other morbidity markers

We found no significant association between current *S. mansoni* infection and PPF, PVD, PSL, anaemia, diarrhoea, vomiting or nausea (Figure 2). This was also the case for all self-reported symptoms (Figure S4), except for blood in stool and having a rash which where both significantly associated with a positive *S. mansoni* diagnosis. For blood in stool this relationship was a significant when including POC-CCA ≥3 (OR=4.20, p=0.003), but not significant when only using Kato-Katz for diagnosing *S. mansoni* infection (OR=1.82, p=0.070) (Figure 2). A similar relationship was observed with having a rash in the last month, POC-CCA ≥3 (OR=2.32, p=0.021) and Kato-Katz (OR=1.79, p=0.049). Current malaria infection was a significant predictor of PVD, anaemia and vomiting (OR=1.20, p=0.011; OR=2.66, p=0.006; OR=2.01, p=0.003, respectively). We also found that PSAC were statistically more likely to have PVD, anaemia and diarrhoea than adults; and SAC were statistically more likely to have blood in stool and vomiting. Males were statistically more likely to have PVD and less likely to have nausea. Individuals with PSL were more likely to have a current malaria infection and be in the PSAC age category, than those without PSL, but this relationship was not significant (p=0.08 and 0.09 respectively) (Figure 2).

### Current infection intensity can predict anaemia and fibrosis but only at very highintensities

As mean intensity of infection increases, we found that the probability of having a positive IP score also increased (B-F) (OR=1.61, p=0.015). We then sequentially removed the high intensity infection and refitted the model and found that this significant relationship was driven by EPGs of >853. Testing positive for malaria was not a significant predictor of IP (B-F) (Figure 3A). Similarly, as mean intensity of infection increased so did the probability of a positive anaemia score (OR=1.40, p=0.037), however, as was seen in IP score this relationship was found to be driven by a very high EPG, >1232. When including both malaria and age class as predictors in the model, malaria was found to be the only significant predictor of anaemia (OR=3.78, p=0.015); while PSAC was a positive predictor of anaemia, this was not significant (Figure 3B).

**Figure 3:**
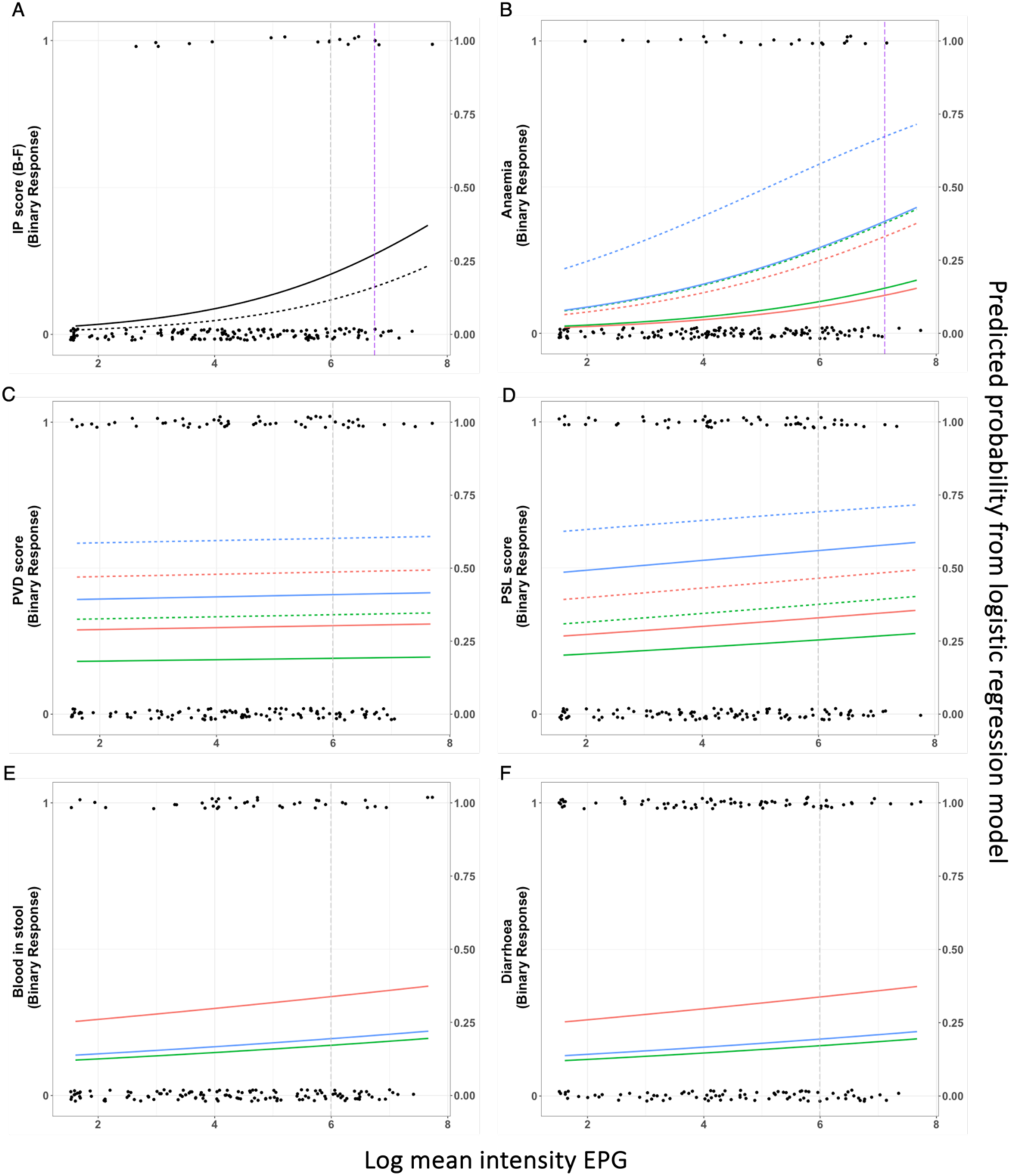
The relationship between current <u>Schistosoma mansoni</u> infection intensity and morbidity. Log mean intensity of eggs per gram (EPG) on x axis, left hand Y axis is binary raw data, right hand Y axis is the predicted probability from logistic regression models, indicated with lines. Colours are age class: blue=preschool age children (PSAC), red = school age children (SAC) and green = adults. The short dashed horizontal lines in the plots represent the predicted probability of morbidity for individuals who are positive for malaria, as estimated by the logistic regression models. The vertical grey dashed line indicates 400 EPG and therefore to the right of this denotes heavy infection, while the vertical purple dashed line highlights when mean intensity of infection in the model significantly predicts morbidity (at this burden and all higher burdens). Image pattern [IP] score measuring periportal fibrosis (A), anaemia (B), portal vein dilation [PVD] (C), left liver lobe enlargement as measured by parasternal line [PSL] (D), self-reported symptoms of blood in stool (E), and diarrhoea (F).

In Bugoto, mean intensity of *S. mansoni* infection did not predict PVD, however malaria infection (OR=2.18, p=0.027) and being PSAC were both positive predictors of PVD, with the latter effect being non significant (OR=2.93, p=0.064) (Figure 3C). PSL was not predicted by increasing mean *S. mansoni* intensity nor malaria infection, however PSAC had a significant association with positivity (OR=3.74, p=0.020) (Figure 3D). SAC had a significant likelihood of having blood in stool and diarrhoea, but there was no difference in the mean intensity of *S. mansoni* infection (Figure 3E and F). No other significant relationships were observed to be predicting the self-reported symptoms, although nausea (OR=2.05, p=0.05) and vomiting (OR=2.07, p=0.06) did have a positive association with a positive malaria infection this was not significant. We found no evidence of a significant interaction term between being malaria and *S. mansoni* infection positive in any of the models.

### Uninfected individuals from Bugoto had higher PVD and PSL than individuals from a non-endemic region

The average PSL and PVD measurements were significantly higher in individuals with undetectable *S. mansoni* infection (negative Kato-Katz and POC-CCA G≥3) from Bugoto compared to the simulated data from the Niamey protocol healthy comparators (Table 2 and 3, Figure S5). These differences were significant apart from PVD in the 80-100cm height category (t -1.4, df 9, p=0.193; Table 2).

**Table 2:**
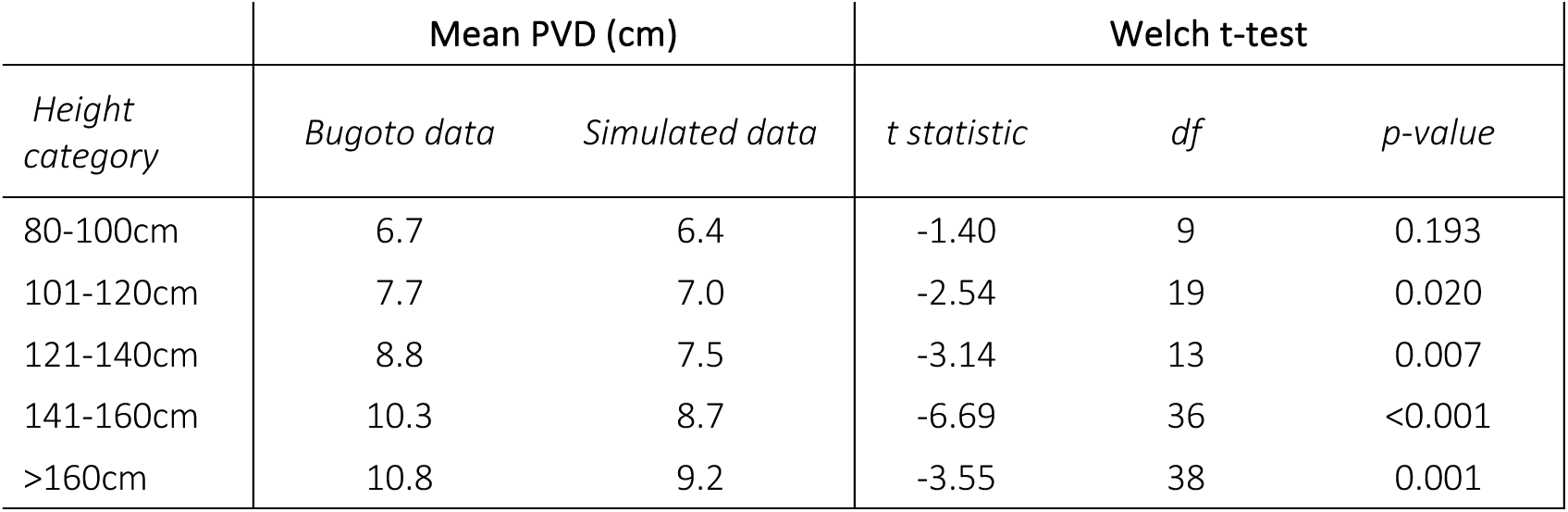
Portal vein dilation (PVD) measurements (centimetres (cm)) in individuals with undetectable *Schistosoma mansoni* infection from Bugoto, Uganda compared with a simulated dataset created from Niamey protocol summary statistics of healthy comparators.

| Height category | Mean PVD (cm) |  | Welch t-test |  |  |
| --- | --- | --- | --- | --- | --- |
|  | Bugoto data | Simulated data | t statistic | df | p-value |
| 80-100cm | 6.7 | 6.4 | -1.40 | 9 | 0.193 |
| 101-120cm | 7.7 | 7.0 | -2.54 | 19 | 0.020 |
| 121-140cm | 8.8 | 7.5 | -3.14 | 13 | 0.007 |
| 141-160cm | 10.3 | 8.7 | -6.69 | 36 | <0.001 |
| >160cm | 10.8 | 9.2 | -3.55 | 38 | 0.001 |

**Table 3.**
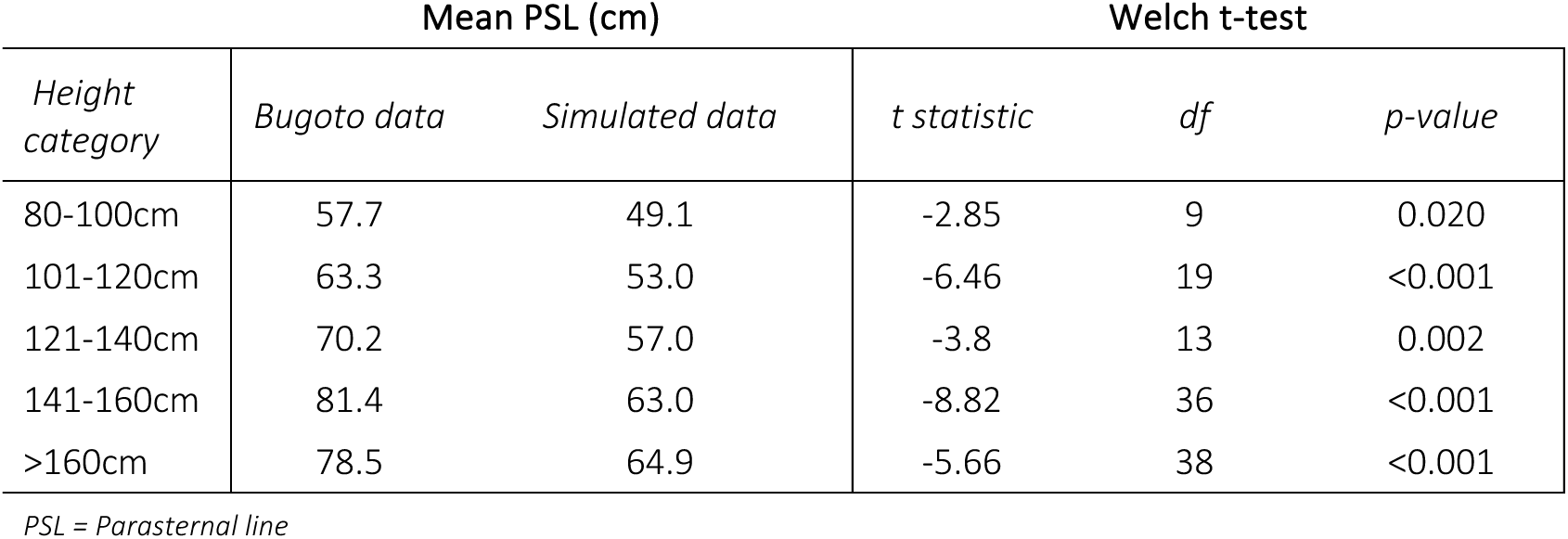
Left parasternal line (PSL) measurements (centimetres (cm)) in individuals with undetectable *Schistosoma mansoni* infection from Bugoto, Uganda compared with a simulated data set created from Niamey protocol summary statistics of healthy comparators.

| Height category | Mean PSL (cm) |  | Welch t-test |  |  |
| --- | --- | --- | --- | --- | --- |
|  | Bugoto data | Simulated data | t statistic | df | p-value |
| 80-100cm | 57.7 | 49.1 | -2.85 | 9 | 0.020 |
| 101-120cm | 63.3 | 53.0 | -6.46 | 19 | <0.001 |
| 121-140cm | 70.2 | 57.0 | -3.8 | 13 | 0.002 |
| 141-160cm | 81.4 | 63.0 | -8.82 | 36 | <0.001 |
| >160cm | 78.5 | 64.9 | -5.66 | 38 | <0.001 |

PSL = Parasternal line

Bugoto PVD and PSL measurements were within the normal range for liver morbidity based on the Niamey Protocol reference graphs [Annex C: Organometry [14]], (Figure 4 – green). However, the confidence intervals extended into the moderate abnormal range (Figure 4 – yellow) for lower height categories in PVD (80-140 cm) (Figure 4A) and multiple height categories in PSL (80-100 cm and 121-160 cm) (Figure 4B) and all points were higher than the simulated means for PVD and PSL from a non-endemic population.

**Figure 4.**
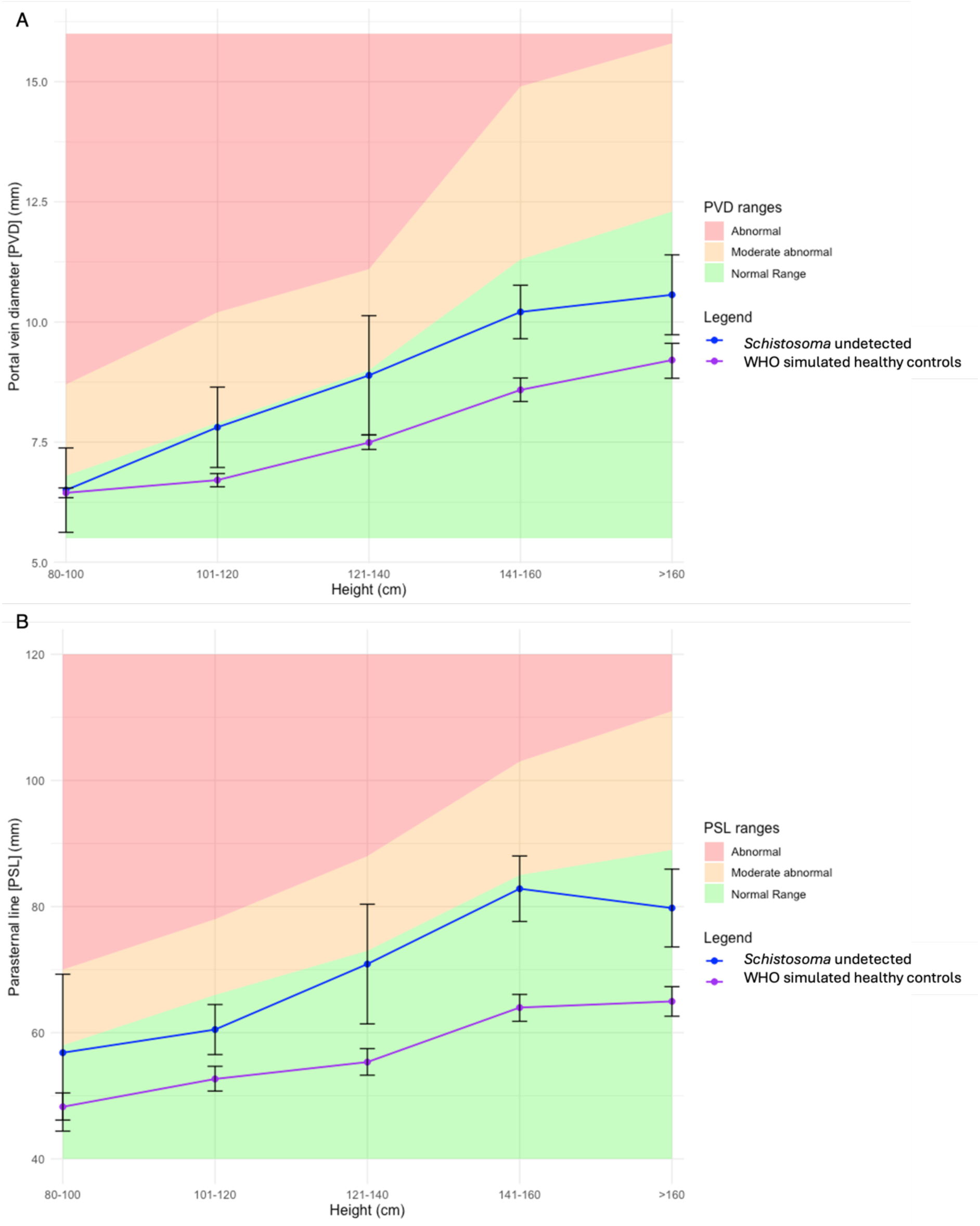
Liver morbidity in <u>Schistosoma mansoni</u> uninfected individuals from Bugoto compared with comparators data from S. mansoni non-endemic areas. The colour PSL ranges are based on the Naimey protocol [14]: normal range (green), moderate abnormal (yellow) and abnormal (red) measurements for (A) portal vein dilation and (B) left parasternal line. Purple line is the simulated data using 1000 bootstraps with sample size matching for height category with the Bugoto study data, and the mean and standard deviation from the Niamey protocol.

## Discussion

In this study we found extremely high *S. mansoni* prevalence levels in Bugoto, Uganda, with 100% of 11–14-year-olds and 73% of the overall population infected with *S. mansoni* (based on Kato-Katz and/or POC-CCA G-score ≥3 diagnostics). We observed relatively low infection intensities (average community = 139 EPG) and low prevalence of a key marker of *S. mansoni*-associated liver morbidity, PPF (IP(B-F) = 9%; IP(C-F) = 4%). However, counter to our expectations, we observed a moderate prevalence of other morbidity markers including PVD (34%), left liver lobe hepatomegaly - PSL (33%) and anaemia (13%) in this community.

Of particular interest, and in stark contrast to our hypothesis that the highest morbidity would be observed in adults due to cumulative infections, we found that preschool age children (PSAC) were most likely to exhibit morbidity, including PVD, PSL and anaemia (50%, 48% and 24% respectively). A recent study which focused purely on PSAC in another highly schistosomiasis-endemic region of Uganda observed similar results: where 8% of the PSAC living in communities near Lake Albert had PPF morbidity (IP (B-F score); [30]); comparable to the 5% we observed in the Bugoto suggesting a potential commonality in early-age morbidity. However, we found much higher levels of other morbidity markers (PVD and PSL), in contrast to Lake Albert community PSAC who had no apparent signs of PVD and only 0.6% had enlarged PSL. Previous studies have demonstrated differing morbidity profiles between these two locations in older age children and adults [26,49,50], but not in PSAC. It is possible that these differences in PSAC’s livers could be impacted by differences in methodology and and/or concurrent malaria infection prevalences, described below. As we were interested in evidence of subtle morbidity, we used a binary system of observed morbidity with PSL or PVD measurements at least 2 SD over the mean of Senegalese healthy comparators, giving a score for either “enlarged” (PSL) or “dilated” (PVD) [14]. In contrast, the study by Pach and colleagues in communities on the shores of Lake Albert used the more stringent 4 SD and over cut-off for either “much enlarged” (PSL) and “marked dilation” (PVD) [30]. If we look at our results using the same stringent cut-offs, we will also observe no PVD in PSAC but would have 7% PSL. The lower PSL observed in PSAC in Lake Albert is likely attributed to the lower malaria infection prevalence, which was 16% in comparison to 32% in our study population and although a current malaria infection was not a significant predictor of enlarged PSL in our study, with higher prevalence in general previous exposure is likely higher in our study location. Despite differences in our methods and findings in this age group, both studies indicate the presence of incipient morbidity resulting from early infections and add further evidence to the need for exploration of the patterns of morbidity and morbidity risk factors among PSAC in diverse geographical regions.

An image pattern of B when measuring for PPF is often not measured due to its indeterminate nature [14], but was observed in our study almost exclusively in the 3–14-year-olds. A similar study which took place in Uganda from 2003-2005 also found only cases of PPF with an image pattern of B in SAC, which they suggested could indicate early changes which, without treatment, would have progressed to more severe fibrosis [40]. This finding almost 20 years prior to our own study highlights that this is not a new phenomenon, but with the relative success of two decades of MDAs, particularly in SAC, potentially resulting in the lower incidence of severe morbidities traditionally found in older age groups (as were also found in our population, albeit at extremely low levels), the focus of studies are changing and looking at these more subtle clinical manifestations which could characterise emerging morbidity in younger age groups.

The age associated distribution we observed could be linked to the timing of MDAs and the impact of sustained treatment in the face of rapid reinfection over many years. This study took place in May 2022 prior to any MDA treatments that year. In Bugoto, annual MDA of SAC began in 2003 [28,40], extending to the whole community (excluding PSAC) from 2004 and became biannual in 2019, but still only in SAC and adults [37]. Consequently, PSAC within this study cohort would not have been part of MDA, while SAC would have experienced between 1 and 12 MDAs based on their age, and adults could have undergone a maximum of 22 MDAs throughout their lifetime if they had lived in this region since the control programme started, although MDA coverage in Bugoto has been very low, especially in adults [51]. Furthermore, those born before the initiation of praziquantel MDAs would have experienced a prolonged period without treatment. This may explain the observed dip in *S. mansoni* specific PPF among 15–20-year-olds. Whilst malaria control programmes which predated the schistosomiasis MDA programme may explain the dip in less specific liver morbidity in 11–30-year-olds (Figure 1 C&D). Regular treatment could reduce the impact of disease in these age groups compared to younger children who are likely infected very early in life and did not receive sufficient MDA treatment and older individuals with pre-existing damage. Our results using the less specific liver morbidities with age as a predictor of *S. mansoni* infection supports this, indicating a loss of the protective effect of treatment in adults over 32 years of age, who were already 13 when MDAs began, sufficient time for lasting irreversible damage to have occurred, compounded by the fact that MDA was mainly administered through schools, and there are only primary schools, and not secondary schools within these communities. Additionally, many individuals in this cohort, particularly those aged 15–20, would have been leaving school completely around the time that MDA programmes were first initiated. While the whole community was included in MDAs from 2004 onwards, individuals who leave school are less likely to receive treatment in subsequent rounds [51], with praziquantel coverage much lower out of school settings in general, both of which would contribute to the increased morbidity observed in these older age groups.

Praziquantel treatment, particularly in children, can significantly reduce early fibrosis and other liver morbidities [22–24,40,40,52–54]. For example, a longitudinal study of SAC, observed a pre-treatment prevalence of 39% PPF and 18% PVD, which reduced to 9.4% and 2.2% respectively in the same individuals one year after treatment [40]. However, many studies have found that in adults or individuals with severe fibrosis, resolution of liver damage post treatment is not as effective [22,24,52]. Furthermore, it is common to see fibrosis (IP C-F) in older populations [27,55]. In a study of 5-90 year olds across three districts in Uganda, found an exponential increase in the likelihood of PPF (C-F) from 26-46 years of age [56], consistent with our results that all incidence of IP C-D, apart from one individual, was observed in older age groups. Importantly, our results strengthen the evidence that MDAs can reduce morbidity, but also highlight the importance of early intervention. Starting treatment in school might be too late for some children, as demonstrated by the 8-year-old with extensive fibrosis and the high prevalence of PVD, PSL and anaemia in PSAC.

Importantly, we found no clear association between most of our morbidity markers and patent *S. mansoni* infections. However, having self-reported blood in stool or a rash within the last month was associated with current infection. Previous research on the use of questionnaires as a rapid screening method in SAC also found an association between egg positivity and self-reported symptoms of blood in stool [57]. However, although SAC did report the highest levels of blood in stool, this was not more likely in those with heavy infections, which is in agreement with results from a very large SAC cohort study which also found a positive association, but no difference between the intensity categories [33]. Blood in stool was reported in two PSAC and three adults who were negative by Kato-Katz and had a POC-CCA ≤3, thus likely caused by something other than current *S. mansoni* infection.

According to the WHO thresholds, the Ugandan community examined here does not meet the criteria for EPHP (<1% heavy infections) nor even that of morbidity control (<5% heavy infections), since 11% of the population had heavy infections. Despite this, our analysis revealed no association between morbidity and heavy infection intensities (EPGs), other than for PPF at levels of over ∼850 EPG. Therefore, our findings suggest limited associations between infection and specific liver morbidity, but only at levels far beyond the WHO threshold of ≥400 EPG. We also observed clear subtle morbidities in those with lower intensities of infection, raising questions about the adequacy of these thresholds in reflecting actual morbidity risk and important ethical considerations regarding subtle morbidities and whether these are a health burden to individuals, their families and communities. WHO thresholds may need to be adjusted to ensure that infection related morbidities are not missed in surveys. In contrast there is also the possibility, albeit it unlikely, that these conditions do not significantly affect the daily lives of individuals nor potentially downstream long-term health and quality of life, which raises critical questions about the necessity and frequency of treatment in such cases [43]. To elucidate these complexities, further qualitative and quantitative research is urgently needed to better understand the real-world impact of morbidities in endemic areas, including creating more meaningful thresholds which guide control strategies to be effective and equitable and ensure the correct allocation of resources.

In our analysis comparing the Niamey protocol measurements of healthy controls from a *Schistosoma* and malaria non-endemic region of Senegal to participants from our study with an undetectable infection, we found consistently higher measurements for PSL and PVD in the participants from Bugoto. Although, mean measurements were all within the normal range, confidence intervals crossed into the moderate pathology range especially for the children, and all were higher than the simulated healthy control means. This higher morbidity could indicate that previous infections leave lasting damage or that subclinical and sub-diagnostic infections, or parasite exposures are indeed causing harm. These differences could also be due to inter-site variation [27], or due to the lack of malaria in this Senegal region. Although the Niamey protocol provides organometric measurements for comparison, the recommendation was always to use values from healthy people of the same ethnic group and region [14], however, in practice this is rarely undertaken, and the Senegalese controls provided in the protocol are usually used, for example [29,30,39,49,58–60], often due to difficulties in finding truly uninfected and unexposed controls from these highly endemic regions.

In this study, the community had high levels of malaria infection as measured by RDT, with an overall prevalence of 31%, which was highest in the younger age groups. Current malaria infection was predictive of both PVD and anaemia, and malaria infection prevalence in younger age groups aligns with the age-related morbidity profiles observed with high PVD, PSL and anaemia in this age group, therefore indicating that either undetectable *S. mansoni* infection or a malaria infection could be driving our results. Researchers observed a significant relationship between PSL and both *S. mansoni* infection and malaria infection in PSAC in Rusinga island, Kenya (also in the north eastern area of Lake Victoria) [29], but as the prevalence of PSL did not decrease after praziquantel treatment, they associated this with an increase in malaria prevalence during the study. However, as mentioned previously, treatment with praziquantel does not always reduce S*chistosoma*-associated morbidity, therefore the observed hepatomegaly may not have been exclusively caused by malaria infection in this study. In a large cohort study of SAC in Western Kenya, the authors found that *S. mansoni* infection only contributed to 7% of the observed anaemia cases whereas malaria and coinfections of malaria and *S. mansoni* contributed 23% and 28% respectively [61]. When examining the role of a hookworm infection and morbidity in our study, there was no association between a detected hookworm infection and any of the morbidity markers in our study group, which contrasts with previous findings in the same district in 2013 which found a significant association between anaemia and having a heavy hookworm infection. These differences are likely due to the much higher prevalence of hookworm and anaemia at that earlier time-point, which were 41% and 44% respectively, rather than 14% and 13% observed in our study, and perhaps an indication of successful STH treatment programs which have run alongside *Schistosoma* MDAs since 2003. These studies along with our own findings highlight the difficulty in disentangling the aetiology of morbidities in regions with co-endemicity and the importance of integrated control programs which have overall benefits in reducing parasite burden and shared associated morbidity.

Of note, individuals in our study were deemed *S. mansoni-*positive if egg positive by up to three days of duplicate Kato-Katz or if negative by Kato-Katz and had a POC-CCA G-score ≥3. This method was used based on latent class analyses by Clark and colleagues [45], which found G3 and over had approximately 90% sensitivity for finding positive individuals. In our study PSAC were more likely to have a negative Kato-Katz test but a positive POC-CCA result (Figure S1). This aligns with findings from other studies, suggesting that low-intensity infections, and potentially reduced faecal output, reduce the specificity of the Kato-Katz technique. Consequently, excluding POC-CCA in diagnostics could lead to underestimations of prevalence, particularly in these younger age groups. The fact that individuals reported as negative using both diagnostics are potentially true negatives, combined with the lack of an association between infection and morbidity markers, suggests that any morbidity observed in the PSAC, who were unlikely to have been previously treated, should be attributed to other factors, such as malaria.

## Conclusions

In our study in Bugoto, a community highly endemic for *S. mansoni* on the shores of Lake Victoria, in Uganda, we observed a low prevalence of PPF, and relatively high prevalence of PVD, PSL and anaemia. We also found a lack of an association between a current *S. mansoni* infection and any of our morbidity markers, and we observed measurable morbidity in those with light or moderate infection intensities. These discoveries add to the emerging body of evidence of a lack of association between EPG and morbidity markers, thus highlighting the need for revised WHO morbidity guidelines. Notably, our study’s use of a comprehensive age range and the use of both the Kato-Katz and more sensitive POC-CCA diagnostics provides new insights, revealing that morbidity patterns can vary significantly across age groups and highlights subtle, yet significant, morbidities that may otherwise go undetected. This comprehensive approach further supports the call for updated guidelines that better reflect the complexities of morbidity in schistosomiasis-endemic regions.

A key insight emerged from the age distribution of morbidities, revealing a relatively high morbidity burden in PSAC and a resurgence of morbidity in older age groups. These findings emphasize the intricate nature of morbidity in a schistosomiasis and malaria co-endemic region and highlight the necessity of further research to unravel these complexities for more effective disease control strategies and strengthened health systems. As we contemplate the new guideline of biannual treatment in all regions with over 50% prevalence [13], we anticipate a decline in severe morbidities [35]. Nevertheless, this will bring with it new challenges in addressing and identifying the subtle morbidities that may elude conventional detection, thus ensuring a comprehensive approach to morbidity control and the well-being of affected communities and individuals of all ages.

## Data Availability

All relevant data are within the manuscript and its Supporting Information files

## Acknowledgements

We would like to thank the communities of, and individuals in, Bugoto for participating in this study. We would also like to thank Dr Sergi Alonso, Dr Jessica Clark, Dr Elias Kabbas-Piñango, Raheema Chunara and Thomas Arme for their invaluable work in the field, and Prof. Jonathan Fallowfield and Dr Timothy Kendall for their advice and opinions on liver morbidities. We are grateful to our funders, R.M.L. supported by the Wellcome Trust program (218492/Z/19/Z; under the supervision of A.B.P., J.P.W., and P.H.L.L.), the European Research Council (ERC) Starting Grant Schisto Persist 680088 (to P.H.L.L.), the Engineering and Physical Sciences Research Council, EPSRC reference: EP/T003618/1 (to P.H.L.L.), and the European and Developing Countries Clinical Trials Partnership (EDCTP) RIA2017NIM-1842 (to J.P.W.)

